# Confirmed central olfactory system lesions on brain MRI in COVID-19 patients with anosmia: a case-series

**DOI:** 10.1101/2020.07.08.20148692

**Authors:** Yannick Girardeau, Yoan Gallois, Guillaume De Bonnecaze, Bernard Escudé, Clarisse Lafont, Gilles Chattelier, Mathieu Marx

## Abstract

**Objective:** Anosmia has been listed as a key-symptom associated with the COVID-19 infection. Because it often occurs without any sign of rhinitis, lesions of the central olfactory system have been suspected. To date, however, there is no evidence that anosmia caused by SARS-CoV2 could be the result of brain damage.

**Methods:** We conducted a case-series on 10 consecutive COVID-19 patients who reported anosmia. Each patient prospectively underwent a validated olfactory test (Sniffin’ Sticks test) and a brain MRI. Results Hypersignal intensity lesions of the central olfactory system were found in 3 subjects on 3D T2 FLAIR and 2D T2 High Resolution images with a lesion involving the olfactory bulbs and/or the orbitofrontal cortex. These 3 subjects showed a severe and persistent loss of smell on the olfactory test. Mucosal hyperplasia of the upper nasal cavities was found in two other subjects with significant smell disorders. There was no MRI anomaly in two subjects with good smell restoration.

**Conclusions:** Anomalies of the central olfactory system could be responsible for anosmia in patients with COVID-19 infection. Further studies are needed to assess the impact on long-term functional prognosis of these lesions.

**Key Result:** Central anomalies of the olfactory bulb and cortex could be responsible for anosmia in COVID-19 infection

## Introduction

COVID-19 is currently responsible for an unprecedented global pandemic (1). However, the actual number of cases is most certainly underestimated due to the scarcity of diagnostic tests and the large share of pauci-symptomatic forms (2).

In addition to respiratory symptoms (3), anosmia appears to be frequently associated with the COVID-19 (4–6). Accordingly, on March 22, 2020, the American Academy of Otolaryngology proposed adding anosmia to the list of symptoms associated to COVID-19 (7).

Post infectious anosmia is well known by otolaryngologists (8–10). It is the main cause with rhino-sinus lesions, and traumatic causes (11). However, the exact pathophysiological mechanisms of post-infectious anosmia remain poorly understood (12). It could involve damage to the olfactory epithelium, to the nerves or the olfactory bulb, or even to the central nervous system (12).

The latter hypothesis could be one of the mechanisms for anosmia cases reported in the COVID-19 epidemic. Two recent publications support it (13,14), both relating to a very similar coronavirus, the SARS-CoV-1.

Our hypothesis was that anosmia caused by SARS-CoV2 could be the result of brain damage. A case series study was conducted on 10 consecutive COVID-19 patient with anosmia who prospectively underwent a brain MRI to identify brain lesions.

## Materials and Methods

### Ethics

The study was approved by the institutional review board of YG and GC hospital. Written informed consent was obtained for all study participants. Data generated and analyzed are available upon request from the corresponding author. The authors declare no conflict of interest.

### Subjects and settings

Ten consecutive patients with anosmia were included in this case series study between 2020 March 23 and 2020 April 17. by YG, M.D. Inclusion criteria were: age ≥ 18 years old; complete loss of smell, without rhinitis at the time of inclusion; confirmed COVID-19 by Polymerase Chain Reaction on a nasal sample or a serology. YG collected retrospectively demographical data, exposure, current medications, medical history and clinical symptoms. Each participant underwent prospectively a brain MRI at least two weeks after the onset of anosmia to avoid accidental staff exposure to SARS-CoV-2. The day before the brain MRI, patients were asked to self-assess the loss of smell (SAS) between: 0 “normal smell perception” and 100 “total loss of sense of smell” and excluded patient who reported a full recovery of their olfaction, and tested included patient for olfactory function using an international validated psychophysical procedure, the Sniffin’ Sticks test (15), where patients have to identify the correct smell between four propositions for 12 different smells. According to the results, the age and the sex, patients were classified as anosmic, hyposmic or normosmic.

### Cerebral MRI protocol

The 10 participants were examined on the same 1.5 Tesla MRI system (GE 450 GEM) using a standardized protocol which included:

- 2D T2 weighted high resolution images, 3.2 mm thick, in the coronal plane covering anterior and middle skull base;
- 3D CubeT2 FLAIR high resolution images, 1.4 mm thick, in the sagittal, axial and coronal planes with extension to the whole brain;
- 3D T1 with and without enhancement images, 1mm thick, in the axial plane, covering the whole brain.

### Image analysis

Nasosinusal cavities and the different structures of each olfactory pathway, i.e. the olfactory cleft, bulb, tract and the corresponding cortex were examined by a neuroradiologist (BE> 30 years experience) and an otolaryngologist (MM> 10 years experience) together. An agreement between both examiners was required to retain an anomaly. Readers were blinded from clinical patients’ features.

### Statistical analysis

Scores are reported as numbers; continuous variables are expressed as means and standard deviations; variables with skewed distributions as medians and interquartile ranges; using R version 3.3.3.

## Results

### General description

Two patients were excluded because they reported a full recovery of smell before the olfactory test and the brain MRI. Ten patients were included (median age, 30 [29-33] years; 60% [6/10] women).

**Table 1:** Patients Characteristics.

| Characteristic | Patients (N=10) |
| --- | --- |
| Age median IQR | 30 [29-33] |
| Male sex no./total | 5/10 |
| Smoking History | 5/10 |
| Current smoker | 4/10 |
| Former smoker | 1/10 |
| Electronic cigarette | 1/10 |
| Alcohol consumption | 7/10 |
| Every day | 1/10 |
| Occasionally | 6/10 |
| Never | 3/10 |
| Any drug abuse | 1/10 |
| Never | 9/10 |
| Less than one time/week | 1/10 |
| Taking any medications in the past two weeks of anosmia onset | 5/10 |
| Paracetamol | 3/10 |
| Diclofenac | 1/10 |
| Iron | 1/10 |
| Melatonin | 1/10 |
| Other | 0/10 |
| Allergies | 6/10 |
| Inhaled allergy | 5/10 |
| Drug allergy | 1/10 |
| Food allergy | 1/10 |
| Medical history | 4/10 |
| Rhinosinusitis disorder | 0/10 |
| Asthma | 1/10 |
| Urethritis | 1/10 |
| Drug induced hepatitis | 1/10 |
| Polycystic ovary | 1/10 |
| Colopathy | 1/10 |
| Other | 0/10 |
| Anosmia as the first symptom | 5/10 |
| Phantosmia | 4/10 |
| Parosmia | 3/10 |
| Ageusia | 7/10 |
| Body temperature $\geq 38^{\circ}\text{C}$ | 3/10 |
| Chills | 5/10 |
| Myalgia or arthralgia | 5/10 |
| Rhinorrhea | 2/10 |
| Nasal obstruction | 3/10 |
| Sore throat | 3/10 |
| Cough | 1/10 |
| Shortness of breath | 3/10 |
| Headache | 7/10 |
| Dizziness | 3/10 |
| Focus difficulty | 5/10 |
| Fatigue | 9/10 |
| Eyes pain | 2/10 |
| Conjunctivitis | 0/10 |
| Abdominal pain | 1/10 |
| Diarrhea | 5/10 |
| Loose stools | 2/10 |
| Nausea | 2/10 |
| Vomiting | 1/10 |
| Skin rash | 0/10 |

Four were current smokers. One had an every day alcohol consumption. Five took medications during the two weeks before the onset of anosmia. Six had an history of allergies and 5 to inhaled allergens (pollens and mites). None of them had an history of rhinosinusal disorder or head trauma. Only 1 had an history of asthma. Anosmia was the first symptom for five. Only 1 had cough and 3 had fever.

Fatigue and headache were the most common symptoms recorded in 9 and 7 patients, respectively. All had experienced a total olfactory loss. Four experienced phantosmia and 3 parosmia. The loss of the function was sudden for 8 patients. Seven had also an impairment of their sense of taste. The median time interval between the onset of anosmia and the brain MRI was 28 days (range of 15-32 days).

According to the Sniffin’ Sticks test, 3 patients were considered as normosmic, 5 as hyposmic and 2 as anosmic the day before the brain MRI.

### Images analysis

A signal anomaly of the olfactory bulb was found in two participants. A hypersignal intensity lesion was found on 3D T2 Flair images covering left OB in the first one, associated with another in the ipsilateral basifrontal cortex (figure 1) while a central hypersignal was found within both OBs on 2D T2 HR images in the second (figure 2). Both subjects showed a severe and persistent olfactory loss with scores on the Sniffin’ Sticks test of 10/12 (hyposmia) and 8/12 (anosmia) respectively. The SAS was also significantly altered in these two patients (65/100 and 100/100). Severe headaches and retro-orbital pain were also reported by the first subject. In the third subject, a hypersignal intensity lesion was found in the right orbitofrontal cortex, which includes the olfactory cortex (see figure 3).

**Figure 1.**
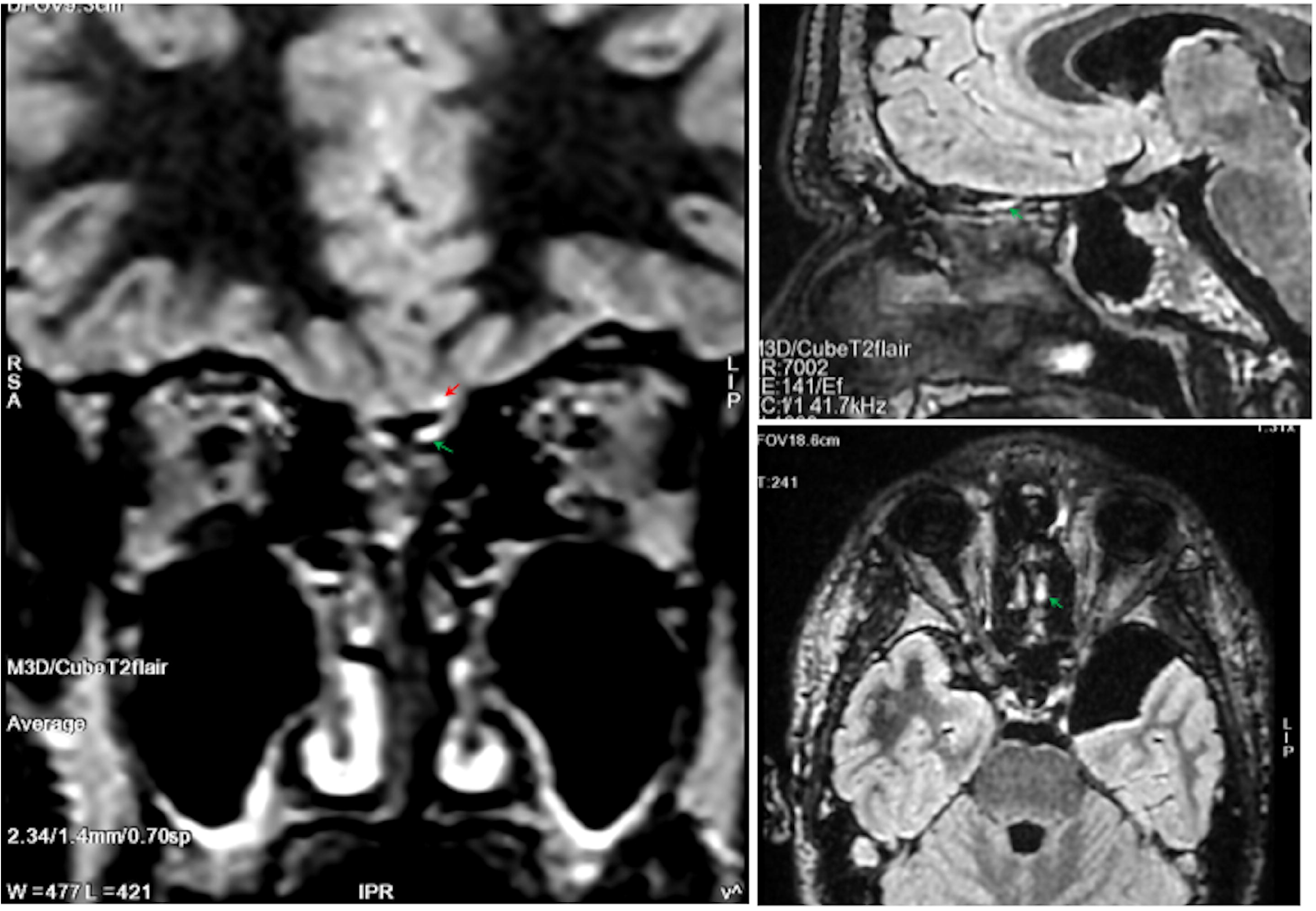
3D T2FLAIR images in the coronal plane (left panel), sagittal plane (upper right panel) and axial plane (lower right panel) showing a hypersignal of the left olfactory bulb (green arrow) and ipsilateral basifrontal cortex (red arrow), in a 21 years old male with persistent anosmia.

**Figure 2.**
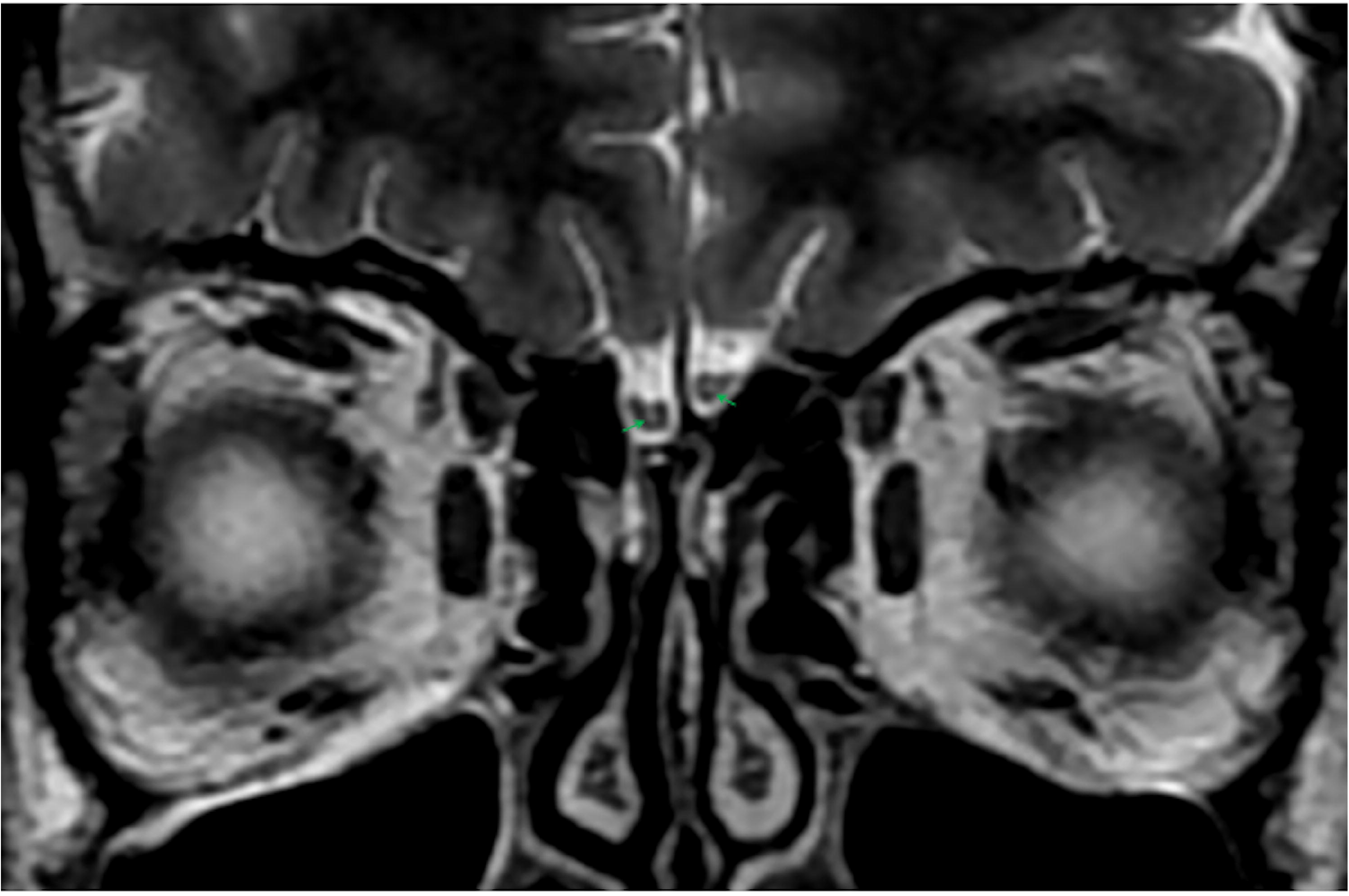
2D T2 High Resolution coronal image showing a bilateral central hypersignal in olfactory bulbs in a 35 years old female with persistent anosmia.

**Figure 3.**
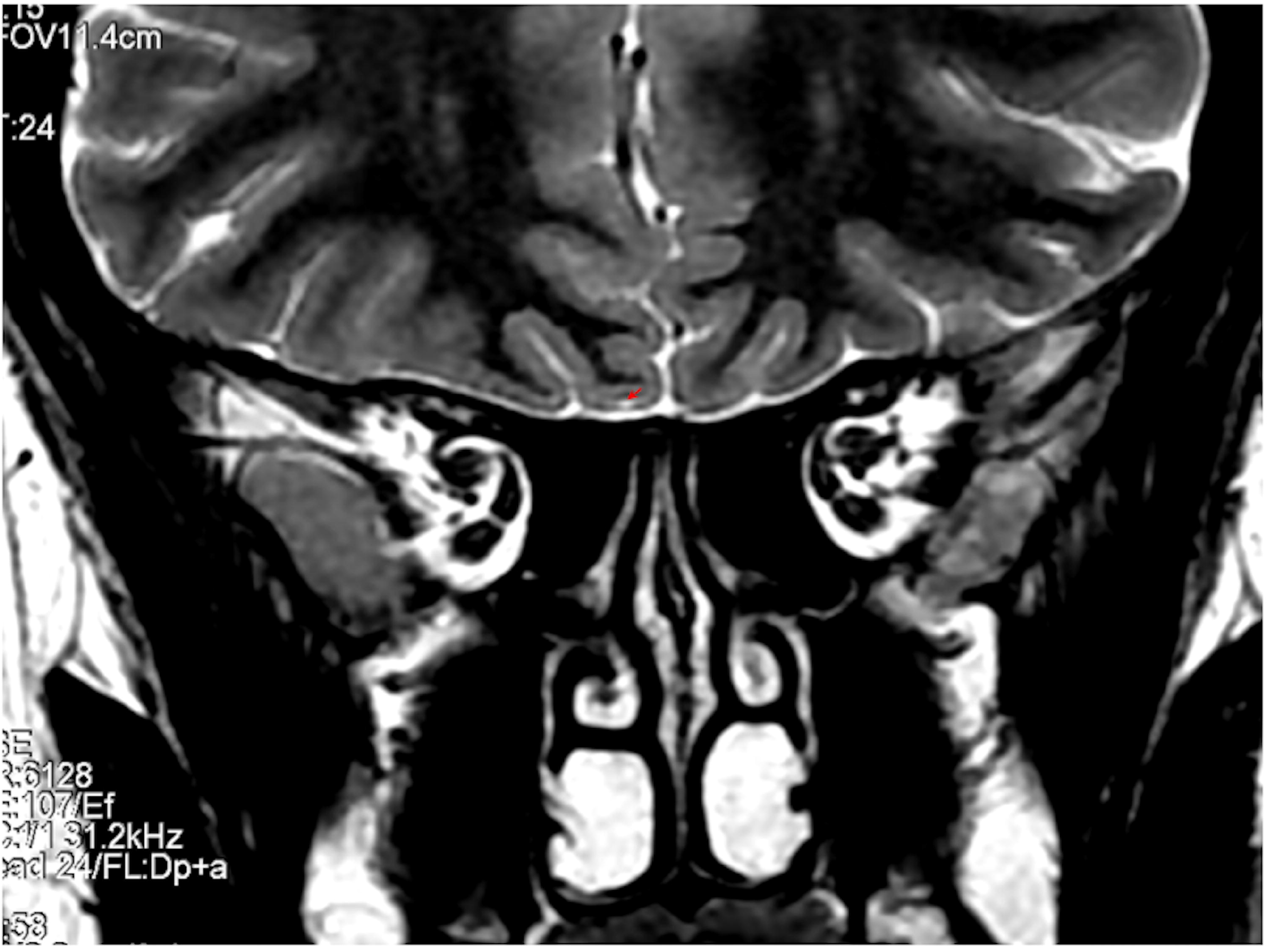
2D T2 High Resolution coronal image showing a hypersignal in the right orbitofrontal cortex in a 29 years old female with persistent anosmia.

Similarly, this participant reported a poor quality of smell (SAS: 100/100), confirmed by the Sniffin’ Sticks test, 5/12 (Anosmia).

Anomalies of the upper nasal cavity were found in two other cases, showing a mucosal hyperplasia of the anterosuperior ethmoidal cells. The quality of smell was also affected in these two subjects with SAS scores of 100/100 and 90/100, and corresponding olfactory performance of 11/12 and 10/12 on the Sniffin’ Sticks test, classified as hyposmia.

Among the five remaining subjects, three showed no anomaly on MRI, reported a good smell restoration with a score of 12/12 on the Sniffin’ Stick test. In the remaining two subjects, the hypothesis of orbitofrontal cortical hypersignal intensity lesions was raised, as in the third subject aforementioned. Unfortunately, the movements during MRI acquisition time precluded any agreement between the examiners. The olfactory scores for these last two subjects were respectively 100/100 and 90/100 for SAS, and 10/12 (hyposmia) for both on the Sniffin’ Stick test.

## Discussion

Several recent studies have identified anosmia as a key-symptom in COVID-19 (4,5). But the mechanisms surrounding anosmia are still under debate (16). Some authors consider that anosmia is the result of the neuro-epithelium inflammation caused by the viral infection (17). But in the context of COVID-19, several patients experienced a total loss of smell without any signs of rhinitis (5).

Therefore, anosmia could result from damage to the central olfactory system. To the best of our knowledge, this is the first case series to report MRI anomalies of the olfactory bulbs and cortex in COVID-19 patients. Our findings suggest that COVID-19 could provoke an inflammation throughout the central olfactory system.

As many other viruses (18,19), coronaviruses could use olfactory tract as a pathway to spread into the central nervous system. Two publications support this hypothesis, both related to a similar coronavirus, the SARS-CoV-1. The first one (13), reports the presence of SARS-CoV-1 in brain samples of victims. The second (14), shows that SARS-CoV-1 was able to penetrate through the olfactory nerves and spread to several brain areas in transgenic mice. Furthermore, SARS-CoV-2 is known to use ACE2 and TMPRSS2 transmembrane proteases for host cell entry and these proteins are both expressed by the olfactory epithelium cells (14,20).

Ideally, brain MRI examinations would have been performed earlier with respect to the onset of anosmia. But the putative exposure to SARS-CoV2 for the radiologic staff for this non-urgent exploration was a real concern. Therefore, most of the brain MRI (8/10) were performed on a dedicated afternoon, more than three weeks after symptoms onset. This delay between the anosmia onset and the brain MRI resulted in the normalization of olfactory performance in 3 patients, considered as normosmic according to the Sniffin’ Sticks test, although their SAS remained unfavorable (25/100 (N= 2) and 10/100 (N=1), 0 “normal smell perception” and 100 “total loss of sense of smell”). Eventually, only patients with confirmed hyposmia or anosmia showed lesions of the olfactory system, whether peripheral or central.

In the same line, several MRI studies on patients with chronic anosmia used 3 rather than 1.5 Tesla MRI to measure the volume of the OBs and to detect subtle signs of cerebral lesions.(21) Restrictions of MRI access due to the epidemic prevented us from using such protocols which might have provided useful data. Still, our protocol was sensitive enough to demonstrate significant anomalies of the central olfactory system in 3/10 subjects, absent in the cases where the olfactory function was restored.

The persistence of severe smell disorders more than 3 weeks after the symptoms onset seems related to central anomalies of the olfactory pathway.

Central anomalies of the olfactory bulb and cortex could be responsible for anosmia in COVID-19 infection. These findings could have immediate consequences for the assessment and the follow-up of patients with anosmia. Further studies are needed to assess the impact on long-term functional prognosis of this identified lesions.

## Data Availability

Data generated and analyzed are available upon request from the corresponding author:
Yannick Girardeau
Hopital Europeen Georges Pompidou - DIH - 20 Rue Leblanc, 75015 Paris

## Acknowledgments

Juliette DJADI-PRAT, Pauline JOUANY, Raphäel HADJADJ, Véronique LAHMAUT, Paul AVILLACH, Anne-Sophie JANNOT.

## Abbreviations

COVID-19: Coronavirus Disease 2019
OB: Olfactory Bulb
SARS-CoV-2: Severe Acute Respiratory Syndrome Coronavirus 2

